# Simulating Emergency Department Boarding Using a Difference Equation

**DOI:** 10.1101/2020.03.19.20039040

**Authors:** Edward G. Brown, Patricia K. Howard, Daniel Moore

## Abstract

**Background:** This paper aims to provide a model that can be used to simulate the effect of patient presentation counts on ED boarder counts and investigate strategies that might be used for managing ED boarding levels.

**Methods:** A boarding simulation model is constructed using a random variable and two regressions that are linked together in a difference equation. The simulation is run under varying constraints, including time interval, presentation counts, and boarder count threshold. Bootstrapping is used to run the simulation a large number of times so that mean and medians can be calculated along with confidence intervals.

**Results:** The method outlined in this paper can be used to simulate the effect of presentation levels on ED boarder counts. Using these methods one can derive quantifiable estimates of time that an emergency department might meet or exceed a particular boarder count threshold.

**Conclusions:** These simulation methods can help an emergency department understand the dynamics of the system in the status quo of normal operations and quantify the relationship of presentation counts and throughput to the hospital. We are hopeful that others may use these methods, adapting, developing, and testing for their own institutions.

## Background

Emergency Department boarding is of particular interest as it affects patient care and safety, operational efficiencies, morale, and revenues. Efforts to understand the dynamics that drive boarding, the primary goal of this paper, are needed in order to minimize these effects. Following is a list of specific problems associated with boarding.

- ED crowding is a symptom of a hospital’s operating at over-capacity, curtailing its ability to absorb the ED workflow. ^1^
- ED boarding has been shown to be a bottleneck in overall hospital flow in multiple studies in the early 2000’s; Creates ED operational inefficiencies in the ED; and Increases mortality of ICU patients. ^2^
- Decreases compliance with sepsis bundles. ^3^
- Associated with delay in antibiotic administration in patients with pneumonia. ^4^
- Poor analgesia with patients in severe pain. ^5^
- Increase in medical errors and in-hospital mortality. ^6^
- Increase in mortality as boarding time increases. ^7^
- ED boarding increases LOS for all patients. ^8^
- Increase in ED hallways - predictive of lower satisfaction scores for the entire hospital stay. ^9^
- Associated with increased LWBS and ambulance diversion leading to a loss in potential revenue. ^10,11^
- Associated with higher ICU occupancy, leading to lower ICU acceptance from the ED, worsening ED boarding, LOS and patient outcomes. ^12^
- Negatively affects the ED staff work environment. It increases job stress, reduces job satisfaction and morale for nursing staff and increases nursing staff turnover. ^13^

## Methods

The following model was developed using two years (2017/2018) of summary data that consisted of patient counts from an electronic health record system used at our (level 1 trauma) academic medical center. The regression equations and bootstrapping were developed using Excel and the statistical software packages SAS and R. Any statistical packages could be used for model definition and development.

IRB officials concluded that our research was not on human subjects as it used only high level aggregate operations data, and as such no IRB review was needed.

The proposed ED boarding model ED uses just three measurements. This approach is taken with the a priori view that hospitals are highly complex systems with a large number of variables (potentially innumerable). For example, for any given patient there are a vast number of measurements which might be taken: acuity, time to see a physician, time to disposition, lab turnaround times, consult times, transport times, and so forth.

Given the high dimensionality of the data which would result from quantifying such a system completely, and the statistical complications of analyzing such data, a much smaller set of measures were chosen. Such an approach is not uncommon when modeling complex systems. ^14^ In a certain sense, the idea is to use a statistical mechanics like approach to dealing with the large and very complex human system.

The three measurements are:

1. Patients boarding at start of day, *a*
2. Patients who start boarding during the day, *b*
3. Patients who stop boarding during the day, *c*

The rationale for using these particular measurements is as follows. Patient boarding is a process which results from inflows and outflows of patients into the ED system which itself connects to two larger systems: 1) the outside world from which patients present and 2) the rest of the hospital system. The boarder count is a result of the interactions of these three systems. Patients who enter this system and are to be admitted will either board or not board. Those that start boarding will increase the count of boarders, and will eventually stop boarding, thereby decreasing the count of boarders. Rather than focus on an hour-by-hour or minute-by-minute calculation to represent this changing total, we use a simple ‘day balance’ total which can be calculated once for each day. In the abstract, it simply represents the net number of boarded patients for a given day.

The ‘day balance’, which will be used as the boarder count for the day for simplicity’s sake, is defined as:

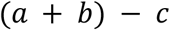

The iteration is defined as:

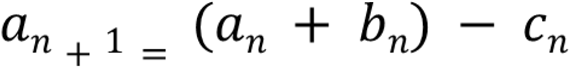

Given these measurements, how might we predict the number of patients who start boarding during the day, *b*? Or the number of patients who stop boarding during the day, *c*?

The first step is to fit two regression models:

1. Patients who start boarding (*y*) as related to Presentations (*x*), for example:

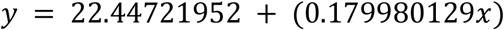
2. Patients who stop boarding (*y*) as related to Patients who start boarding (*x*), for example:

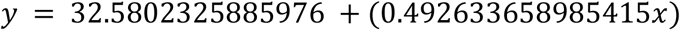

A brief explanation may be useful as to why these regressions were chosen. The first regression may seem self-evident in that presentations are the primary input to a system which will produce discharges, admitted patients, and boarders. To some degree it is natural to look for linkages between new presentations and patients who start boarding in any given day. The second regression was chosen for two reasons. Firstly, there will be natural turnover rate of boarded patients and this regression can represent that rate in the system. The second reason is based on the a priori view that an increased rate of new boarders may put more stress on the system as a whole and a subsequent increased rate of patients who stop boarding would be more likely to follow.

*Note: if no associations can be found for these particular measurements, other associations can be explored. As there is variation between emergency departments, there’s no guarantee that these particular associations will be found. The dynamics of each particular system will dictate which associations you investigate*.

## Results

Using the linear equations we solve for the ‘day balance’ (number of boarders) over a given number of days using the iteration. For example, one can estimate the number of boarders per day from a given set of parameters across 30 days by allowing the number of presentations to vary randomly within realistic upper and lower bounds. These upper and lower bounds can be determined by analyzing your ED’s presentation history. In other words, you can begin the simulation process using real measurements and then control the parameter that you are varying randomly and feeding into your iteration.

Furthermore, one can set a threshold target for number of boarders you do not wish to meet or exceed and display the results of these simulations graphically:

**Figure 1:**
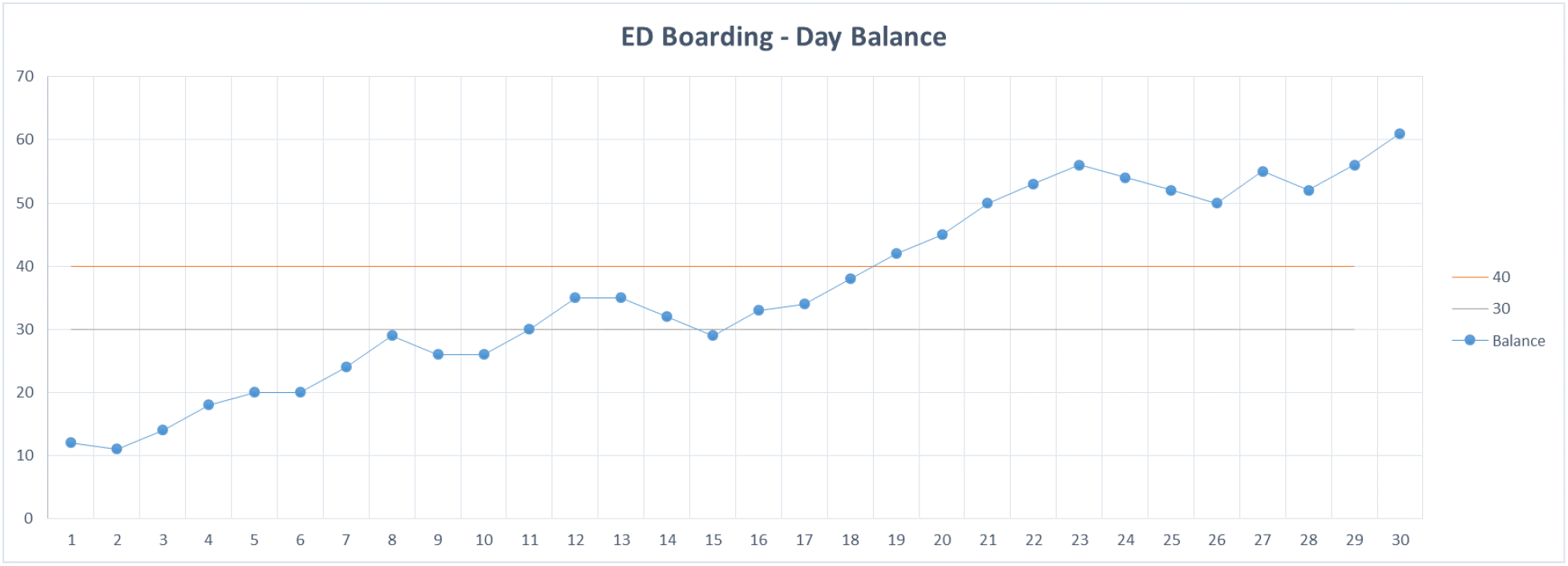
Simulation which crosses threshold.

In this simulation, the threshold of 40 boarders is exceeded continually after day 18. Obviously, every time you refresh the simulation, allowing the number of presentations to vary randomly, the results will look very different. Here is another graphical result of the same simulation method:

**Figure 2:**
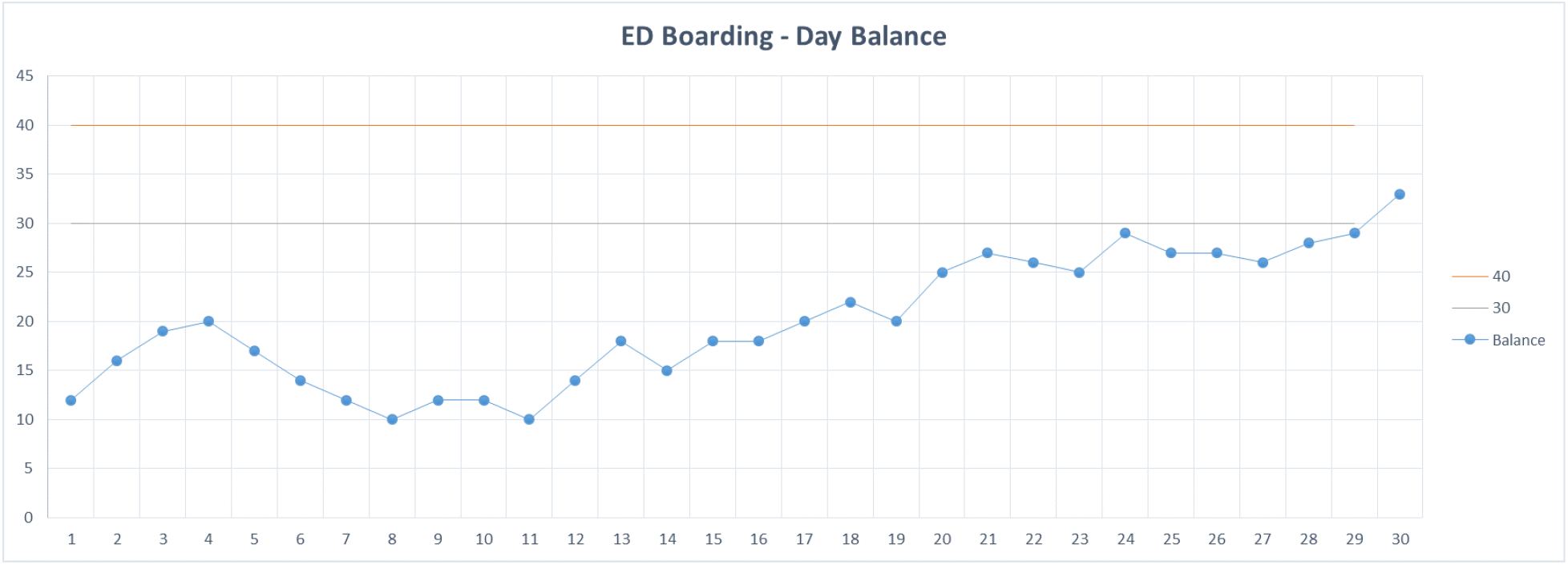
Simulation which doesn’t cross threshold.

In this simulation, the threshold of 40 boarders is never reached.

So how can this simulation be useful if each run yields different results? One way to utilize these simulations is through bootstrapping. By running the simulation set a large number of times, say 10,000 (i.e. 10K 30 day periods), we can store the number of days in each 30-day period of the simulation that our boarder count meets or exceeds our threshold (30 or 40 for example) and calculate a percentage. We then store this percentage for each of the 10K trials, finally calculating the mean and median for the simulation as well as confidence intervals for our estimation. For example, using our iteration with a lower bound of 230 presentations, an upper bound of 290, and a threshold of 40 boarded patients, one run of our bootstrapped simulation reports (M = 0.71, Mdn = 0.73, SD = 0.05) with a 95% CI [0.6,0.8]. Repeated runs of the simulation return similar results. We can plot a histogram of our simulation as well.

**Figure 3:**
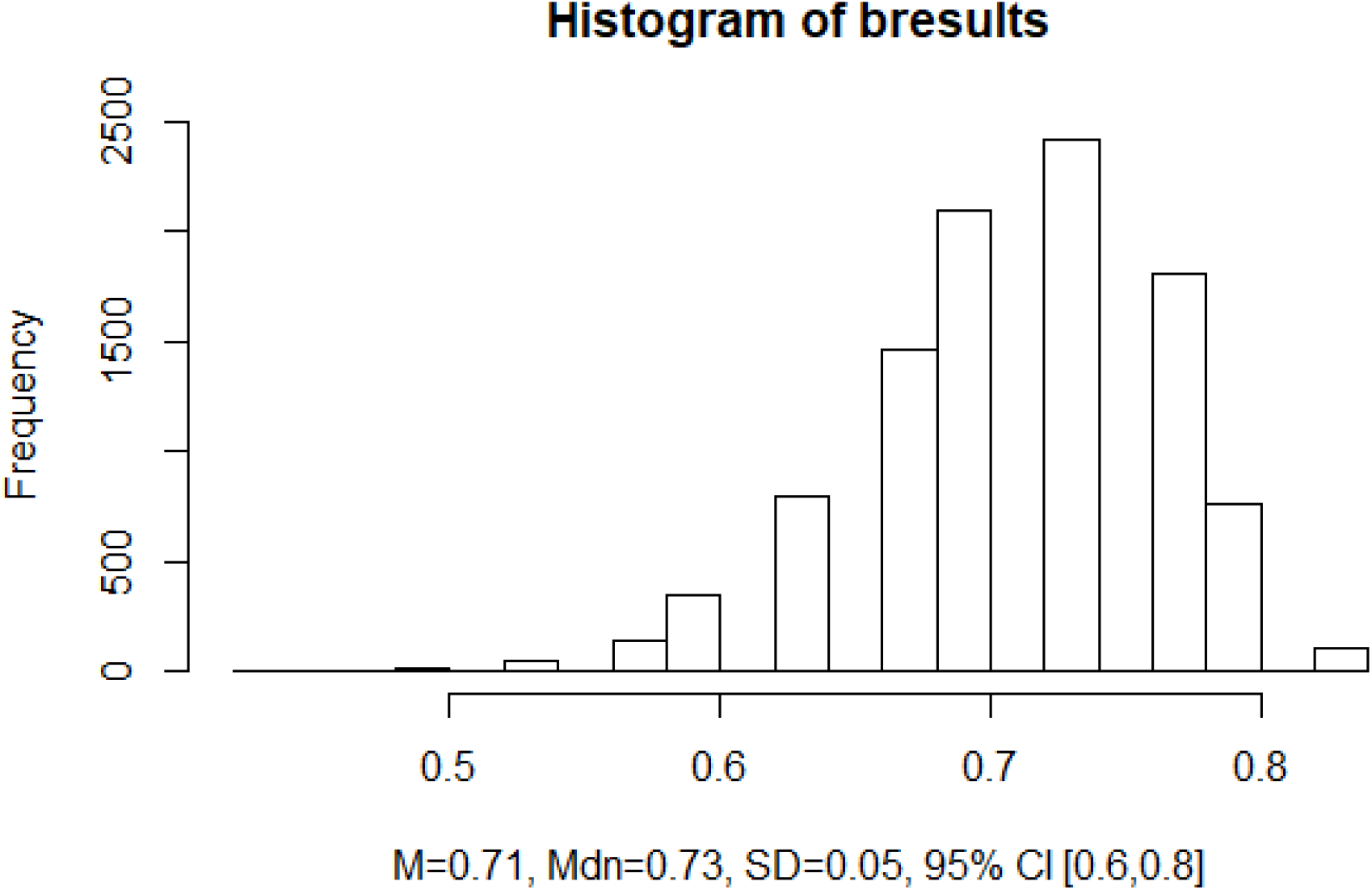
bresults is our bootstrap result set.

## Discussion and Limitations

Assuming that an emergency department has some ability to control the number of presentations during a given day (e.g., going on divert) one can try a couple of different methods to see how setting an upper presentation limit might affect your boarder counts.

The first method is to set a constant upper limit—in other words divert would start as soon as the presentation upper limit is reached and remain on for the rest of the day. The second method is to set a targeted upper limit, which would kick in once a boarder threshold has been reached—in other words divert would start when the threshold is reached and would stop once the boarder count is below the threshold.

Using the simulation method outlined in this paper, one can adjust the lower limit, upper limit, and threshold. Example output is below. The percentages show the percentage of time (using output from the bootstrapped simulations) that the boarder threshold would be met or exceeded.

Example output from method 1 (constant upper limit) reporting median % time at or above threshold:

**Figure 4:**
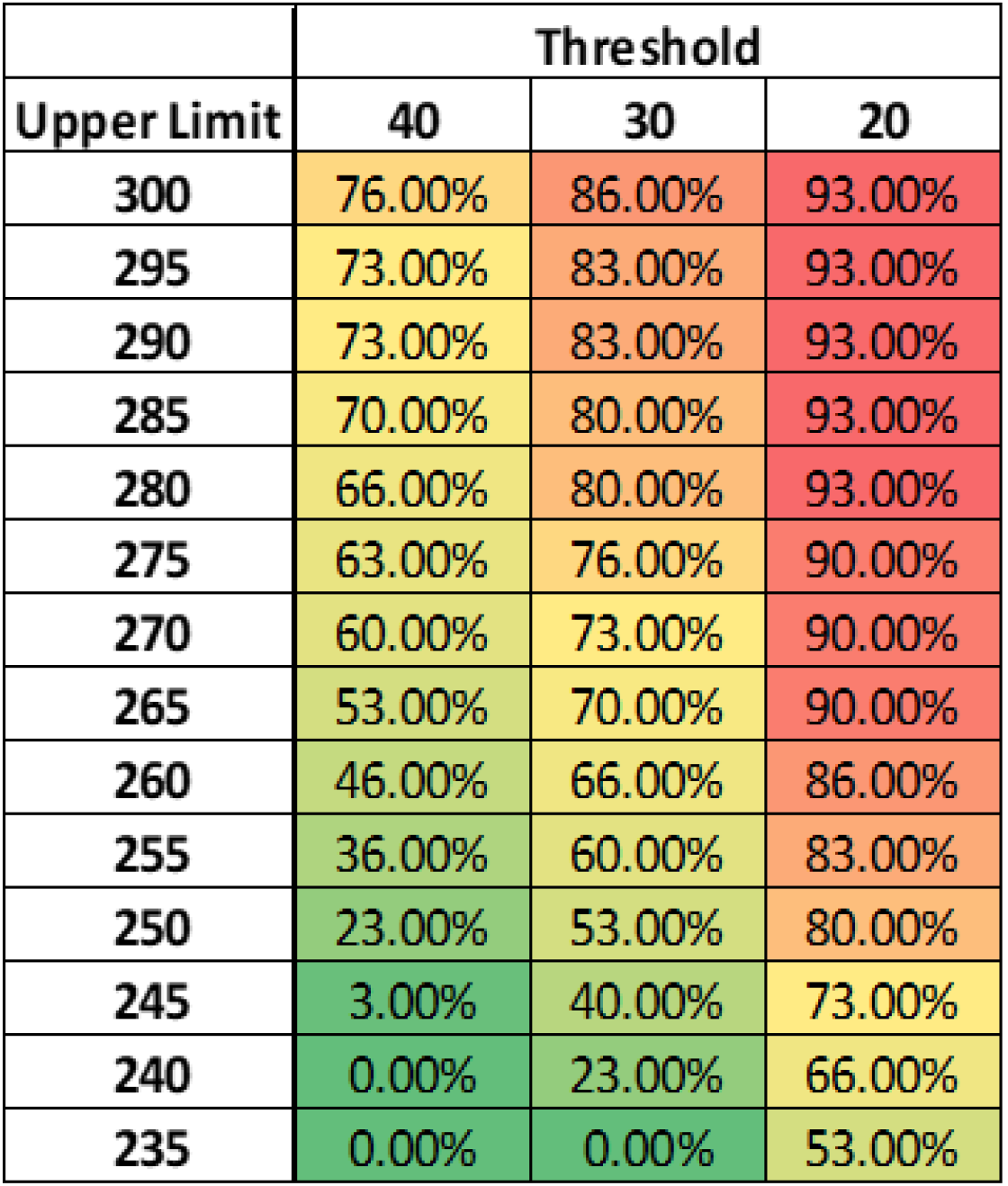
Results of simulation method 1.

*Note that the lower the threshold, the greater the percentage of time that the threshold is exceeded for equivalent upper limits*.

Example output from method 2 (targeted upper limit) reporting median % time at or above threshold:

**Figure 5:**
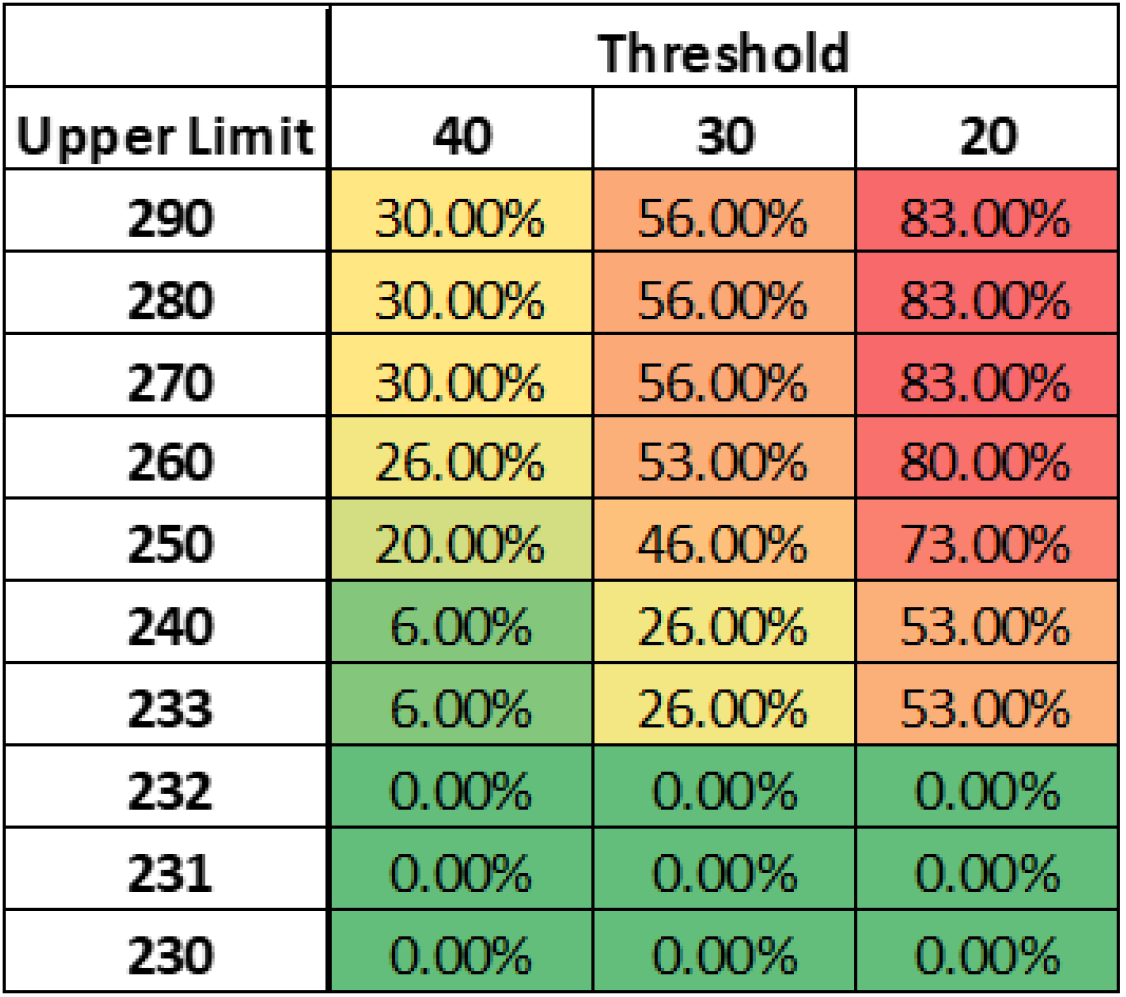
Results of simulation method 2.

*Note that in this method the simulation essentially provides a daily upper limit past which boarding can become problematic, in this case > 232. The simulation allows for finding the optimal upper presentation limit to use once your boarder threshold has been met or exceeded*.

This output is based on the regression equations above and would obviously vary considerably across emergency departments, just as the regression equations themselves would.

In reality, it is unlikely that by going on divert the patient presentation count would drop to zero until lifted. This complication is an important factor to consider. Further model development could account for this limitation by adding a certain percentage of presentations to the model even after the threshold has been reached. Study of divert and the amount of patients that continue to present after divert is triggered would be needed to derive an appropriate number.

It’s also difficult to know to what extent changes occurring in real time will affect the overall system. For example, if throughput of patients moving from the ED to admitting beds is increased for any number of reasons, the boarder count could drop at a rate higher than simulated. Further study of hospital dynamics could investigate more fully, perhaps using differential equations which might better reflect the changing relationship of the two systems.

The method outlined in this paper can be used to simulate the effect of presentation levels on emergency department boarder counts assuming that regression equations can be fit which adequately model the dynamics of the system. Using these methods, while tuning the limits and threshold, one can derive quantifiable estimates of time that an emergency department might meet or exceed a particular boarder count threshold. While variation is to be expected, the use of bootstrapping and the derived confidence intervals can aid in decision making regarding how an emergency department might try to manage presentation counts.

While we chose 30 days for the development of the simulation, the timeframe can be adjusted as needed. In addition, the presentation upper and lower bounds can be adjusted, as can the threshold. It’s important to note that the point of these simulation methods is not to make highly accurate predictions. Rather, the goal is to define a reasonable estimate of a variable percentage of time that an ED may have boarders above a certain threshold given system dynamics and presentation counts.

Finally, keeping in mind the limitations noted above these simulation methods can help an emergency department understand the dynamics of the system in the status quo of normal operations and quantify the relationship of presentation counts and throughput to the hospital. It may ultimately be as useful to simulate and understand how presentation counts push a system to meet a threshold as it is to simulate and understand what might happen once the threshold has been exceeded.

## Data Availability

Code samples are available in a github repository.

https://github.com/edwardbrown/ED_Boarding_Simulation_Difference_Equation

